# Fully Personalised Degenerative Disease Modelling - A Duchenne Muscular Dystrophy Case Study

**DOI:** 10.1101/2022.07.28.22278103

**Authors:** Evan Baker, Peter Challenor, Ian Bamsey, Francesco Muntoni, Adnan Y. Manzur, Krasimira Tsaneva-Atanasova

## Abstract

Predicting the trajectory of rare degenerative diseases can be extremely beneficial, especially when these predictions are personalised to be relevant for a specific patient. These predictions can help inform and advise patients, families, and clinicians about the next stages of treatment and care. Obtaining such predictions, however, can be challenging, especially when data is limited. In particular, it is important that these predictions do not rely too heavily on general trends from the wider afflicted population while not relying exclusively on the, potentially sparse, data from the patient in question. We present a case study, wherein a modelling framework is developed for predicting a patient’s long term trajectory, using a mix of data from the patient of concern and a database of previously observed patients. This framework directly accounts for the temporal structure of a patient’s trajectory, effortlessly handles a large amount of missing data, allows for a wide range of patient progression, and offers a robust quantification of the various uncertainties. We showcase this framework to an example involving Duchenne Muscular Dystrophy, where it provides promising results.

## Introduction

Degenerative diseases cover a wide class of conditions where the state of a patient deteriorates over time. These are often genetic, or have genetic components, and often have no known cure, with only treatments to slow the progression or mitigate the symptoms. Examples include cystic fibrosis ^1^, Huntington’s disease ^2^, Alzheimer’s ^3^, spinal muscular atrophy ^4^, and multiple sclerosis ^5^. These diseases are often not completely understood, but due to their chronic nature, improved treatment and prognosis can have a great impact on a patient’s quality of life. As such, statistical modelling of these diseases can be valuable as it can aid understanding of these diseases, at least with regard to their general trajectory, reaction to treatments, and how different patients experiences can differ. This can be especially valuable for personalised medicine, as data on the patient in question, instead of only data on the wider affected population, can be used to tailor treatments and predictions.

Modelling of these diseases, however, can be challenging. The trajectory of a patient can be complex, and so standard linear assumptions over time can be insufficient. This can be characterised by non-monotonic disease progression, such as biphasic slow (or non-existent) declines in health followed by fast declines. This issue is confounded by the fact that many degenerative diseases are rare, and so data can be limited, which makes it difficult to learn complex relationships from the data. Similarly, the disease progression trajectories are heterogeneous, i.e. the trajectory of one patient with the disease can vary massively to the trajectory of another. For example, one patient could have a relative period of stability followed by a sharp decline, while another patient could deteriorate slowly but consistently. Learning all trajectories, for all patients, can be difficult without an abundance of data on each patient. Furthermore, because the trajectories can be complex in shape, it is desirable to have data in all stages (of interest) of the diseases, however this is often not the case as data can be lacking for some or all patients. For example, perhaps the early symptoms of a disease relate strongly to the final outcome, but data on these early symptoms may very well be limited, with only a few patients ever being observed so early. Alternatively, perhaps a sharp decline exists for most patients after a certain time point, but if no data is available after this time point, most models will struggle to capture this sharp decline.

Many different strategies exist in the wider literature for modelling disease progression. There are several features that need to be directly captured, or controlled for, by such a model. One important feature is time; degenerative diseases progress over time, and so how this relationship is captured is core to any degenerative disease model. Another key feature is inter-patient variability; these trajectories can differ for different patients, and so there needs to be some way of accounting for that. Treatments also need to be captured, as they can have a big impact on the resulting trajectories.

One strategy is to learn a small set of representative trajectories, and then classify patients as belonging to one of these trajectory ‘classes’ ^6,7^. This can be effective, and it can also be intuitive and easily explainable - non-statisticians can be made familiar with the different classes of patients. When classifying a patient, uncertainty can also be provided via probabilities of belonging in each class. Treatments can also have an impact on which class each patient falls into. The downside is that less of the individual variability is captured. If a patient doesn’t fit well into any of the trajectory classes, in general they will still be allocated a class (the one they are most similar to). Similarly, a patient might fit well into one class to begin with, but diverge later. This is especially likely if treatments given to the patient change over time. As such, these clustering methods are well-suited for stratification, that is, describing a disease more generally, and providing simplified (generic) predictions for a given patient, but they can struggle with more precise and personalised modelling goals.

Another option is to perform regression, rather than classification. This requires a carefully planned structure for the various features, and there is a wide range of possible modelling decisions. Time can be explicitly modelled, or time (age) can be included as a covariate to be regressed on. Including time as a covariate can be easier, making use of more of the wider regression methodology. Explicitly modelling time requires a careful choice for the model of time, such as a state-space model or a hidden Markov model, but it ensures a causal directionality. Additionally, when predicting the future, explicitly modelling time helps ensure uncertainty grows over time. Similarly, inter-patient variability can be explicitly modelled (to varying degrees), ignored entirely, or separate models can be made for each patient. Fitting separate models relies on having sufficient data on each patient, and does not provide (or make use of) any population-wide information. It does however ensure each patient receives a fully-personalised model. A fully general model, ignoring patient variability, does not provide the personalised care that patients with chronic disease need. It does however make use of population-wide information, and does not need sufficient data on any specific patient. Modelling inter-patient variability can be the preferred outcome, making use of random effects (also known as partial pooling), wherein parameters are personalised, but a global distribution for them is learnt. However, knowing what random effects to include can be difficult, and depends on the type of model used. Including treatments then depends on the other modelling decisions. If time is included as a covariate, it can be difficult to include treatment effects that change over time, at least not without massively increasing the dimensionality of the problem.

For example, Alzakerin et al. ^8^ used second order autoregressive models (AR(2)) to model Huntington’s disease. This is a model that explicitly models time, but separate models are used for each patient. Evers et al. ^9^ used Dynamic Linear Models (DLMs) to model Parkinson’s. In this case, time is again explicitly modelled, with separate models again for each patient, but certain parameters in the models (the trend between each time step, and the variances) are made the same. Severson et al. ^10^ used hidden Markov models for disease progression, with the disease able to progress between different latent states over time. In this model, these latent states are different for each patient, but the parameters controlling the random processes are the same between patients, with the exception of an additive inter-patient random-effect term and a personalised parameter for the effect of treatments. This type of model does assume there are discrete states of health, which may not always be reasonable.

Other models regress on time (or age), providing a functional form for the trajectory. This form needs to be chosen well to ensure the relevant relationships are captured. Hamuro et al. ^11^ outlined several simple functional forms for Duchenne Muscular Dystrophy (DMD). Some of these are polynomial, while others are made up of pre-defined piecewise linear regressions. Some of these models include random effect parameters. Simple functional forms like this can be data-efficient, but overly simplistic forms rely heavily on prior-expertise and are limited in scope. Schulam and Saria ^12^ used a combination of simple parametric and spline terms for hierarchically modelling population, sub-population, and individual trajectories. Gaussian Processes are then also used for each patient, providing structured uncertainty that grows further away from data, imitating the effect one would get from explicitly modelling time. This is a promising strategy, and whilst an explicit model for time might be preferable, especially if predictions of the future are desired, it does show that the alternative, when done well, can also be useful.

Another strategy is to make use of differential equation models, using data to estimate the various parameters. Lennie et al. ^13^ made use of an ordinary differential equation for DMD, with some of the parameters allowed to vary for different patients. Monte Carlo Markov Chain is then used to estimate the various parameters. This idea relates to Uncertainty Quantification (UQ), where a complex numerical model for a system (in this case, a degenerative disease) can be developed, making use of knowledge of the underlying processes involved. Statistical methodologies can then be used to estimate model parameters, accounting for the various uncertainties Such practice is common in many fields of science, including climate ^14^, engineering ^15^, epidemiology ^16^, and indeed healthcare ^17^. This is perhaps the gold standard for modelling, but it can require an incredible amount of understanding about the underlying processes, and because of the complexities of these processes (and thus models), estimating the various parameters can become challenging ^18^.

Another relevant technique is deep learning models (such as neural networks). These can be used in many disease modelling strategies, and can be beneficial because of the complex relationships involved ^19^. However, such methods usually require a relative abundance of data to perform well, and with rare diseases this is not always the case. Similarly, the lack of well-calibrated uncertainty from these models can be concerning, especially with regard to degenerative disease care.

In this paper, we outline a DMD case study which serves to exemplify a modelling framework that we have developed. Our modelling framework is based on DLMs and takes some of the best attributes from various modelling approaches. This includes explicitly modelling time, which allows for a patient’s treatment to change over time and allows for comprehensive uncertainty estimates for future prediction. We provide personalised models for each patient, while still drawing information between patients via extensive use of partial pooling. We have enough complexity to capture the various relationships, but the methodology is also simple enough to be explainable and easily generalised. We can also model multiple clinical metrics at once, incorporating the structure between these, which is not always (easily) possible with some trajectory modelling approaches (especially those methods which do not make use of latent disease states). Additionally, because our approach is based on DLMs, we separate observational noise from structural variability, allowing more complex trajectories than our parametrisation would usually allow, and we can easily accommodate missing observations.

A description of our modelling framework is given in Section. Afterwards, this framework is applied to an example involving DMD in Section, before finishing with concluding remarks in Section.

## Methodology

In our modelling framework, observations are modelled as

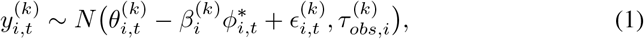

with *i* representing the individual patient, *t* representing time, and *k* indexing the clinical outcome of interest. This treats the observations as coming from several distinct latent state variables. Together, these latent state variables control what the ‘true’ value is for the observation. The standard deviation, 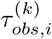 accounts for the presence of observational noise and random fluctuations around the latent ‘true’ value. With this, the ‘true’ value progresses over time, and we occasionally take measurements, which we observe with some error. Missing data does not prevent the existence of the latent states, it only means that there is no observation of 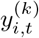 for a given *i, t*, and *k*. Whilst we assume normality for the observations here, this need not be the case more generally, and other distributions can be used instead.

The first latent state, 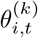, represents the healthy state of the patient. Over time, this can progress (for example, because the physical abilities of the patient in question improve as they reach maturity). This latent state broadly controls how the patient would perform, were they healthy and did not have the degenerative disease. Mathematically, we model 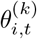 as

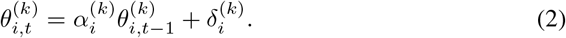

Here, 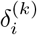 controls how much the patient improves by over time, but 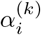 allows this improvement to level-off over time (as the patient reaches maturity).

The next latent state, 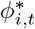, represents the progression of the degenerative disease.

This latent state is subjected to a positive constraint transform, specifically the softplus transform, which is common in many machine learning tasks:

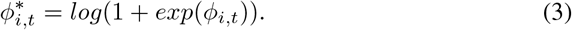

This transform ensures that the effect from the disease is always of the same sign (and so can be forced to only negatively affect the patient).

The underlying *ϕ*_*i,t*_ disease state then progresses as:

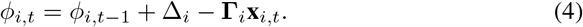

*ϕ*_*i,t*_ progresses linearly over time at a rate of Δ_*i*_, although, due to the way it interacts with the non-linear healthy state, this can still result in more complex declines in patient wellbeing being captured. Similarly, because of the softplus transform, the negative effects from the disease can have a delayed onset, as negative values of *ϕ*_*i,t*_ only result in 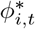 getting increasingly close to zero, and so a very negative value for *ϕ*_*i*,0_ corresponds to a long lag before any (measurable) effects from the disease manifest. **Γ**_*i*_**x**_*i,t*_ is a simple regression term representing the impact of various interventions and treatments. **x**_*i,t*_ are the actual treatments administered. These can be binary or categorical variables, representing which treatments are administered, or continuous variables (such as how much of a drug is given). **Γ**_*i*_ then captures a patient’s responsiveness to a specific treatment. With **Γ**_*i*_**x**_*i,t*_, the treatments can slow the progression of the disease, and the treatments can change over time.

The final latent component is then 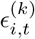

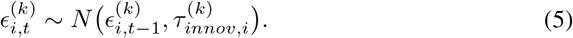

This is simply a random walk term, which captures any localised trajectory changes a patient might experience. In this way, each patient is allowed to progress in a more complex way than the equations above would allow, and said complexity would not be assumed to be observational error. It might be expected that we include these random ‘innovations’ in the equations for both *θ*_*i,t*_ and *ϕ*_*i,t*_, but we find in practice that this can cause identifiability issues between the two.

The above equations outline a personalised model for any given patient, for any amount of time, for any amount of clinical outcomes. However, there is not always enough data on each given patient to allow such a complex models to be fit. This requires us to make use of data from multiple patients in the prediction for any given patient. However, we specifically do not want to make global, population-based predictions; we are still interested in personalised predictions. It was with this in mind that the above equations were designed, and to tackle this issue we make heavy usage of random effects, or ‘partial-pooling’. Each of the parameters outlined above controls a specific property of a given patient, and so, because there is still an overarching pattern of trajectories amongst patients with the same degenerative disease, these parameters should be similar between different patients. This similarity may be very weak, or it may be very strong, but importantly this allows us to capture *some* information contained within the wider-population’s data that is relevant for individualised predictions.

As such, we aim to learn what the population-wide distribution is for each of the outlined parameters, which facilitates the estimation of patient-specific parameters.

For example, 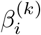, which is the one parameter not yet discussed, controls the scaling for the disease state for each observed clinical outcome. If a deteriorating patient resulted in an increasing metric for one clinical outcome, but a decreasing metric for another, then the disease state would need to be given a different sign for each. 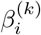 allows for this, along with varying magnitudes of impact.

This scaling parameter, 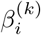, which controls the general relationship between different clinical outcomes, should be very similar for all patients. If a deteriorating disease state is related to a declining lung capacity in one patient, then we would expect this same relationship to hold for a different patient (to some degree). With this in mind, we aim to learn what the overall average value for this parameter is, and how much this parameter varies across the population:

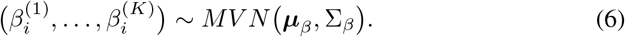

Here, we jointly model all the 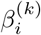 values for the *i*^*th*^ patient with a multivariate normal distribution. If the diagonal entries of Σ_*β*_ are very small, then all patients have a very similar value for the 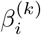 values (approximately ***μ***_*β*_). If the diagonal entries of Σ_*β*_ are very large, then each patient has a very different value for 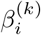. This allows us to give each patient their own value for 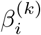, even with a limited number of data points for each patient, because some information about 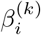 can be shared between patients. Additionally, by using a multivariate normal distribution, we capture correlations between the 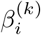 values for different clinical outcomes. We would expect the impact of the disease to have a similar effect on the different clinical outcomes, and so it is sensible to have the outcome-specific parameters be correlated as well.

Note that the first 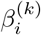 (when k = 1) can be set to 1, without any required estimation, as the scaling of *ϕ*_*i,t*_ itself renders it redundant.

The same approach can be applied to other parameters within the model. With 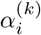, we want to constrain the value to within (0, 1). To do so, we make use of the logistic transform, which is often used in logistic regression to convert continuous numbers into probabilities.

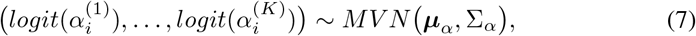

We constrain 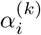 to be between 0 and 1 because negative values cause oscillations in the patient’s natural clinical outcome, and greater than 1 causes the clinical outcome to naturally increase exponentially. These attributes may be possible in other situations, but in this work we do not consider it realistic.

To improve identifiability, we also add constraints to the rate of disease progression. We constrain Δ_*i*_ to be positive, which forces the disease state to naturally increase over time, representing the progression of the disease. This constraint is not completely essential, as if the disease state instead decreased over time, then the 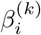 coefficients could flip sign to accommodate this. Of course, as is, we have to carefully choose which clinical outcome is the first outcome (*k* = 1), because if that outcome does not have a respective 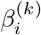 value, then the disease state must have a negative impact on that outcomes values. This should not be difficult, but it is an important point to remember. For the positive constraint, we make use of the softplus transform.

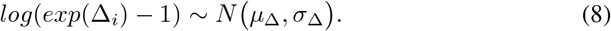

Note that this parameter is modelled with a univariate normal distribution, as it is global across all clinical outcomes.

We also add constraints to the treatment effects **Γ**_*i*_. When the amount of data is low, and the variety of treatment plans in the data set is low, then it can be easier for a model to simply learn that increased treatments are correlated with worse patient outcomes. This obviously is the incorrect direction of causality, and is not desirable. The explicit time structure of our model should avoid this, but with observational data (rather than experimental data), where the variety of treatment plans and the density of data can both be low, it can still be possible for the model to struggle. Constraining the treatment effects to be non-negative, whilst crude, forces the model to learn the more interesting structures present in the other latent states. This results in the following equations for **Γ**_*i,p*_, where p indexes the individual treatment effects.

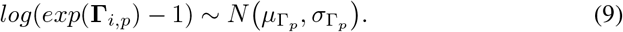

One parameter where it would not be sensible to perform partial-pooling (at least not directly) is 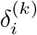. Whilst 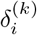 does control for a specific property of a given patient (the rate of natural improvement the patient would otherwise experience), the actual numerical values for 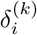 are somewhat meaningless. This is because there is a strong interaction with the parameter 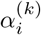, which controls the shape of the patient’s natural progression. A large value for 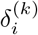 would usually mean very fast natural change in the patient, but if 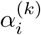 were 0, then it would actually result in no natural change over time for the patient. As such, without knowledge of 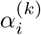, the value of 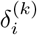 does not have any intuitive interpretation, and the values of 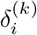 for different patients may not have any similarities. A solution to this, is to instead relate the values of 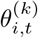 at a certain time. If, for example, we were to choose some sufficiently late enough time, where all patients would be expected to be fully mature, then this value has an interpretation, and this value should have some interpretable correlation between patients. We can then use this, as well as the value of 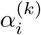 to back-infer what the value of 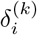 is.

In other words, we say:

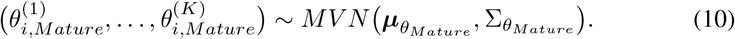

We can then see how 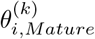 relates to 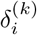 by simply iterating Equation (2):

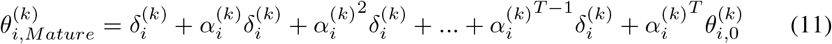

where T is the number of timesteps needed to reach maturity (this will need some clinical insight, in this work we assume age 20 for T). We can then see that this is in fact a geometric series (plus an additional term):

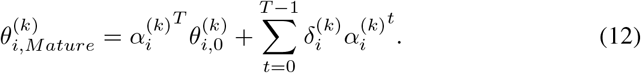

Which then has a known analytic formula:

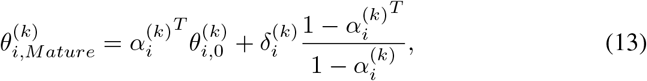

and so we can obtain the value for 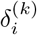:

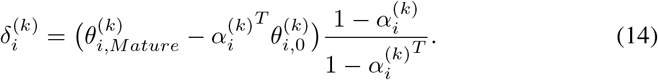

The observational and innovation standard deviations, 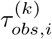 and 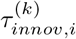 are both modelled using truncated normal distributions (truncated to be positive):

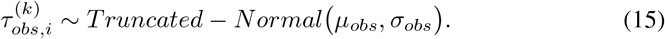

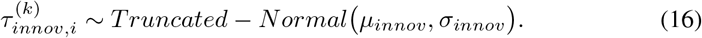

To finish the model framework, we then also have to estimate the initial values for the latent states, for all patients:

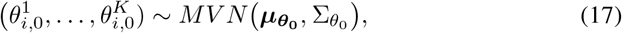

The initial disease state has a similar issue as 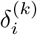, in that the numerical value for *ϕ*_*i*,0_ can be less meaningful, although this issue is not as severe as it is for 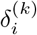. If the initial disease state is negative, this represents a partial delay for the onset of symptoms. The length of this delay, however, depends on the rate of disease decay, Δ_*i*_. As such, we choose to model the time when the disease state *ϕ*_*i,t*_ would reach zero in the absence of any treatments, 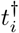, rather than *ϕ*_*i*,0_ directly (which, with the softplus transform, is when the *gradient* of 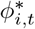 would reach 0.5):

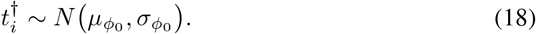

We can then easily get the value for *ϕ*_*i*,0_ from 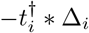 (because 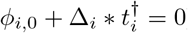). Here we can see that it’s not too difficult to conceptualise what *ϕ*_*i*,0_ represents, but 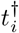 is slightly more interpretable when there is a delay to the symptoms’ onset, and so we choose to mode, this instead.

Collectively, this outlines the general framework for modelling individual patient trajectories. Specific changes can also be made where clinical knowledge is available, and where specific clinical outcomes or treatment effects require it. From here, given a data set, the various hyperparameters in the system can be estimated (either via maximising the likelihood, or via MCMC, or otherwise), providing a model that can produce individualised predictions for the patients, and a model that also contains an understanding of the global patterns amongst the patients.

### Application - DMD

To showcase this modelling framework, we consider an example involving Duchenne Muscular Dystrophy (DMD) ^20^. DMD is a degenerative disease which causes muscles to weaken, leading to progressive walking difficulties, loss of deambulation, respiratory insufficiency, among other symptoms. The disease predominantly affects boys, and results in very early death, around the ages of 20-30. This disease is genetic, and there is no-known cure, although treatments do exist ^21^.

The progression of the disease can vary wildly between different patients ^7^. This increases the difficulty in providing support for those affected, not only as it can impact the optimal treatment plan, but because general guidance for family is hindered. For example, when a wheelchair may be required can be an important moment in the care of the affected, and preparing for such a moment can be taxing, both logistically and emotionally. This large degree of variation can also complicate clinical trial planning and interpretation

With a wide range of potential trajectories in the population, being able to provide personalised predictions for the health of a specific patient can be valuable, both for clinicians, and those who have to live with the condition.

We have access to a database of anonymised clinical records of patients with DMD, via the NorthStar Clinical Network Database (https://www.northstardmd.com). The dataset contains over a thousand patients, although the quality and quantity of data on each patient varies considerably.

Included within this dataset are several gradings of a patient’s ability to perform certain tasks, graded on a three-point scale. These tasks include the patient’s ability to walk, stand on one leg, climb a box, and other such tasks. Collectively, these can be combined into an overall assessment of the patient’s motor skills, called the North Star Ambulatory Assessment score (NSAA), which varies from 0 to 34^22^. This is a very valuable resource, as it is a single metric which summarises a patient’s function, and is common enough to result in a widespread data set, including as a primary endpoint in clinical trials. We also have access to other information about patients, including age, height, weight, the various treatments they are on, and some other measurements from the patients.

With NSAA as our primary measurement, we can use the model outlined in section to capture the population-wide trends, but we can also obtain patient-specific models, and therefore patient specific predictions, for NSAA. Note that, technically, NSAA is a discrete variable, only allowing whole numbers between 0 and 34. This would suggest that a normal distribution for NSAA may not be the perfect choice, and instead a binomial distribution, might be a better choice. Such a modelling choice is entirely possible with this framework, as the latent states 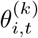 and *ϕ*_*i,t*_ could instead represent the latent binomial probability of a success, and no further changes would be needed. In practice, we find the normal distribution sufficient for this example, providing better confidence for predictions where the NSAA is not near 0 or 34, and the effects of rounding predictions to the nearest valid whole number do not appear particularly problematic.

Figure 1 shows these NSAA trajectories for a selection of patients. From these, we can see that, whilst there is a general trend that many patients follow, there is also a large variety in patient experience.

**Figure 1.**
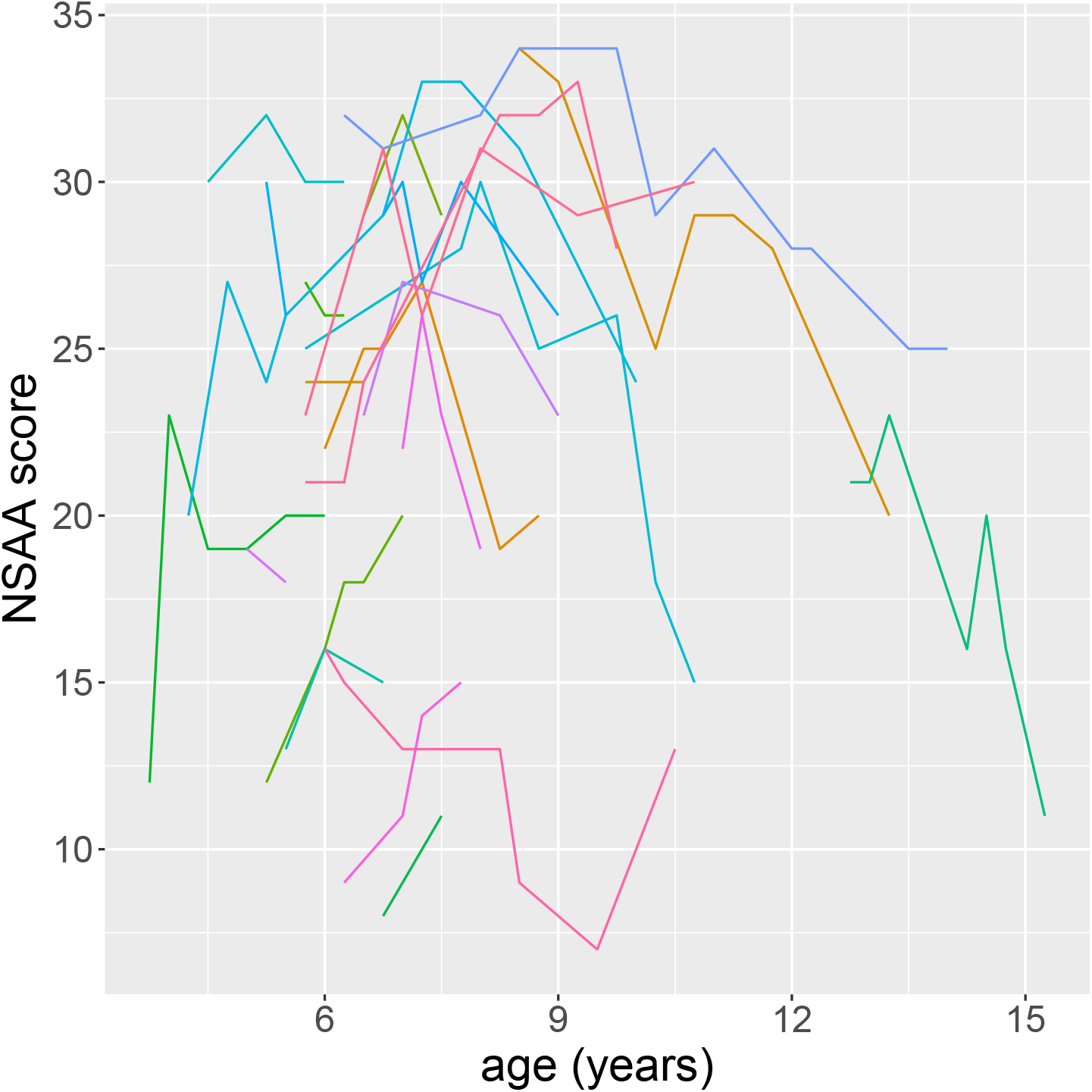
A plot showing several trajectories of NSAA for different patients.

We also make use of other clinical outcomes. One is walk time, which measures (in seconds) the time taken to walk/run 10 meters. Another is rise from floor time, which measures the time taken to stand up straight from a lying down position. We have slightly less data on walk time and rise from floor time, but they are a still a valuable and abundant source of information (4867 total entries for walk time, 4636 for rise from floor time, and 5989 for NSAA).

For treatment effects, we have access to the steroid regime the patients are on. This includes the steroid schedule, which we use as our treatment effect (as its prescription is a standard of care recommendation). The data also includes how much steroids the patient is on, and which steroid, but we do not make use of this here for this example, as most patients in our dataset follow the same steroid dosage regime (see Figure 3 in Birnkrant et al. ^23^). The steroid treatment can potentially change over time (which is not a problem, as **x**_*i,t*_ is explicitly allowed to vary over time), although the steroid schedule does not often change over time in the dataset. The steroid schedule is not always explicitly recorded at all timesteps, however, a patient is unlikely to change their steroid regime without clinical oversight, and so we can safely assume that a missing entry here means that no change has occurred. The type of steroid is a categorical variable, which can be extended into multiple binary columns of **x**_*i,t*_. Some patients have records for a dosage of steroids, but it is never recorded which schedule they were allocated. In these cases, we still make use of these patients but allocate the steroid type “unknown”. In total, we have 5 regimes the patient can be on: “None”, “daily”, “intermittent 10.10” (10 days of steroids, then a 10 day break), “other”, and “unknown”. “None” is the default, and so the effect of all other regimes are parametrised as the difference from “None”.

We have data recorded from the patients roughly every 6 months. However, this fluctuates, as the availability of clinic slots is not always at exact 6-month intervals. Additionally, these 6-months are not in sync between patients. For example, one patient might have their first appointment 1 month after their 4^*th*^ birthday and roughly every 6 months afterwards, but another might have their first appointment 3 months after their 4^*th*^ birthday (and roughly every 6 months afterwards). Collectively, this influences how we choose the time step for our model. We might initially want to use a 6-month time step, but the issues just mentioned make this somewhat imprecise. The perfect time step might instead then be 1-day, but this would be computationally intractable with this number of patients. We decided on a 3-month time step, which allows the time steps to roughly follow the every-6-months that the clinic appointments aim for, but with an additional level of precision indicating whether each data point is early during a 6-month window, or late. Additionally, while the aim might have been to have one clinic appointment every 6-months, from the moment of diagnosis to the moment it stops being practical, in practice there are lots of missing appointments. This is not a problem with our modelling framework, as the latent ‘true’ NSAA values (and walk time values, etc.) continue to exist, they are simply unobserved for some time steps. The age of diagnosis also varies for each patient, and so when data on each patient begins to appear also varies. This is also not a problem, as, given a fixed start time for the model, this simply means there are often no observations for patients early on. This is similarly the case for observations late in a patient’s progression. Here, we choose the patient at age 3 as the initial time for each patient, and we stop considering observations after age 20. We could extend this to age 0 and age 30, but the amount of observations outside the 3-20 range is very small (only 25 NSAA observations, out of 5989, are outside this range), but the computational burden of modelling an extra 13 years would be very high.

Before providing this data to the model, we transform the clinical outcome data to enforce certain, known, constraints. For example, the NSAA data must be an integer between 0 and 34, and so we use the transform *logit*((*x* + 0.5)/35), which then scales the interval [−0.5, 34.5] to (−∞, ∞). As such, model predictions after reversing the transform, and rounding the results, will produce integer values between 0 and 34. Similarly, walk time must be positive, and so we use the softplus transform, which scales the interval from (0, ∞) to (−∞, ∞). After transforming the data, the observational data is standardised so that it has mean of 0 and standard deviation of 1, and the input **x**_*i,t*_ values are scaled to have a minimum value of 0 and a maximum value of 1.

Within this work, we choose to estimate the hyperparameters via MCMC. Performing inference in a Bayesian setting provides a full quantification of the various uncertainties. This is valuable in this setting, as there is a lot of uncertainty regarding the various parameters, especially on a patient-specific level, with a limited amount of data for each patient. This does mean we have to provide prior distributions for the various hyperparameters. This can be a great benefit, as it allows us to incorporate clinical knowledge. This is especially useful for the parameters surrounding the healthy state, as we do not have access to healthy patients in this example. Unless stated otherwise, our priors are reported with values relevant to the standardised scale.

For the multivariate covariance matrices, we separate the correlation matrix from individual standard deviation terms ^24^. The correlation matrices are then given LKJ distributed priors ^25^, with a shape parameter value of 1. This provides a uniform distribution over the possible correlation matrices.

On the unstandardised scale, the mature state mean for NSAA is given a normal distributed prior, itself with a mean of 34 on the untransformed scale (so it has a mean of *logit*((34 + 0.5)/35), and standard deviation of 0.5, and the mature state standard deviation is also given a truncated (above 0) normal prior with mean 0 and a standard deviation of 0.5. This is quite a strong prior, but because of how NSAA is set up, a score of 34 should be expected for almost any healthy adult. Also on the unstandardised scale, the mature walk time state mean is given a normal distributed prior with a mean of 2 on the untransformed scale, and a standard deviation of 1; the mature walk time standard deviation is also given a truncated normal prior with mean 0 and a standard deviation of 1. The same priors are used for the rise from floor time mature states. Similar prior distributions are given for the initial states. The initial NSAA state mean has a prior mean of 15 on the untransformed scale, with a standard deviation of 1, and the initial NSAA state standard deviation has a prior mean of 0 with a standard deviation of 1. The initial walk time state mean has a prior mean of 7 on the untransformed scale, with a standard deviation of 1, and the initial walk time state standard deviation has a prior mean of 0 with a standard deviation of 1. The same priors are used for the initial rise from floor time states.

The other parameters in the model are given priors on the standardised scale. 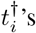 mean and standard deviation are both given *N* (20, 5) priors (with the standard deviation prior truncated to be above 0). Treatment covariates are given *N* (−8, 2) priors for their mean, which assumes the treatments have little-to-no effect but does allows the alternate possibility, and truncated *N* (0, 1) priors are used for their standard deviation. The innovation mean and standard deviations are both given truncated *N* (0, 0.1) priors, as are the observational error mean and standard deviations. These allow for a small range of values around zero they could take, as we believe most of the variation in the data is *not* a result of random noise.

The mean for the healthy patient shape, 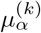, is given a *N* (0, 3) prior, which coupled with the logit transform, provides a fairly uniform prior over the values of 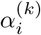 between 0 and 1. All other mean parameters are given weakly informative *N* (0, 1) prior, and all other standard deviation parameters are given a truncated *N* (0, 1) priors.

## Results

To fit this model, we split the data into training and testing data. The training data is used to fit the model, learning the posterior distributions for the various parameters, and the testing data is not given to the model, and therefore can be used to check the quality of the model’s predictions. To split the dataset into training and testing data, we randomly choose 200 patients. For these 200 patients, a random amount of clinical appointment data is held-aside, varying from 80 *−* 20% (removing the most recent appointment data). This provides a wide range of representative potential scenarios the model may have to predict in practice, which we can use to assess the performance of the model.

Using the training data, we fit the model via MCMC using nimble ^26,27^. We run two chains of MCMC, use a burn-in of 200, 000 then sample for a further 800, 000 iterations, and a thin of 800.

We can then extract predictions from this model, and Figure 2 shows example NSAA predictions for six patients.

**Figure 2.**
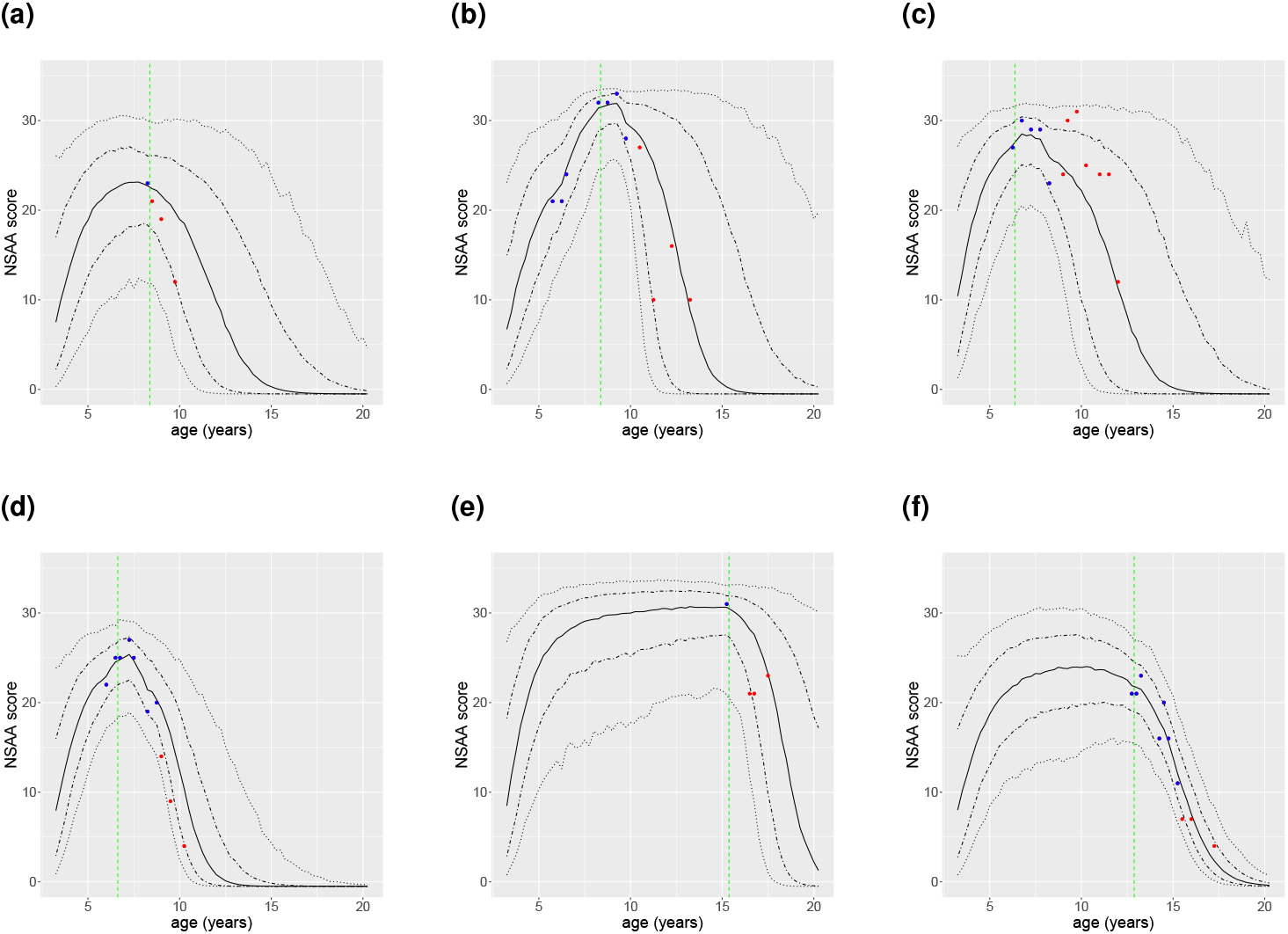
Predictions of NSAA for six patient. Blue dots are measurements that are provided to the model, red dots are measurements that were held-aside and were not given to the model. The central black lines represent the median prediction, with the outer lines representing the 70% and 95% prediction intervals. The horizontal green lines represent when a steroid intervention occurred.

**Figure 3.**
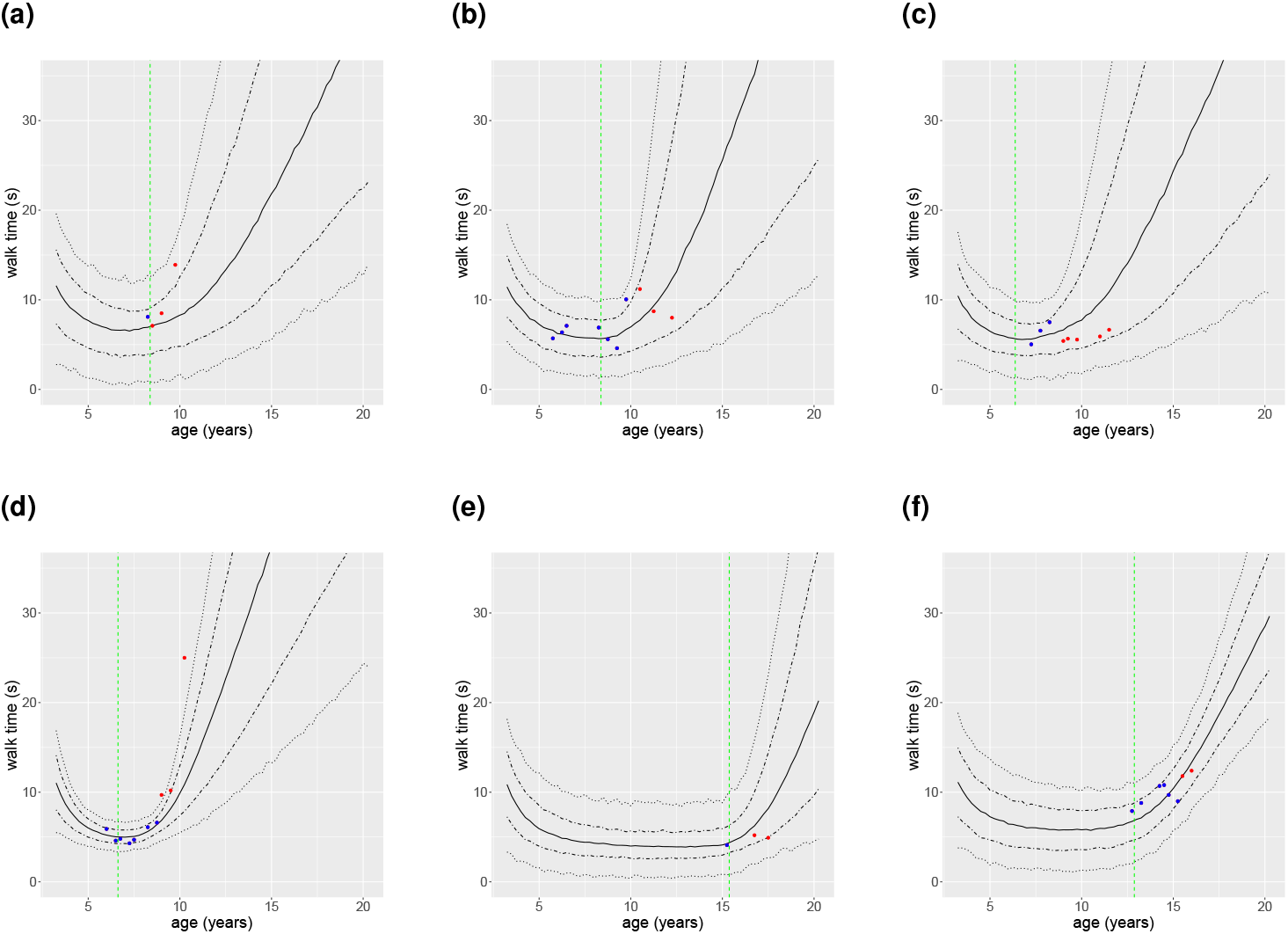
Predictions of walk time for six patient. Blue dots are measurements that are provided to the model, red dots are measurements that were held-aside and were not given to the model. The central black lines represent the median prediction, with the outer lines representing the 70% and 95% prediction intervals. The horizontal green lines represent when a steroid intervention occurred.

**Figure 4.**
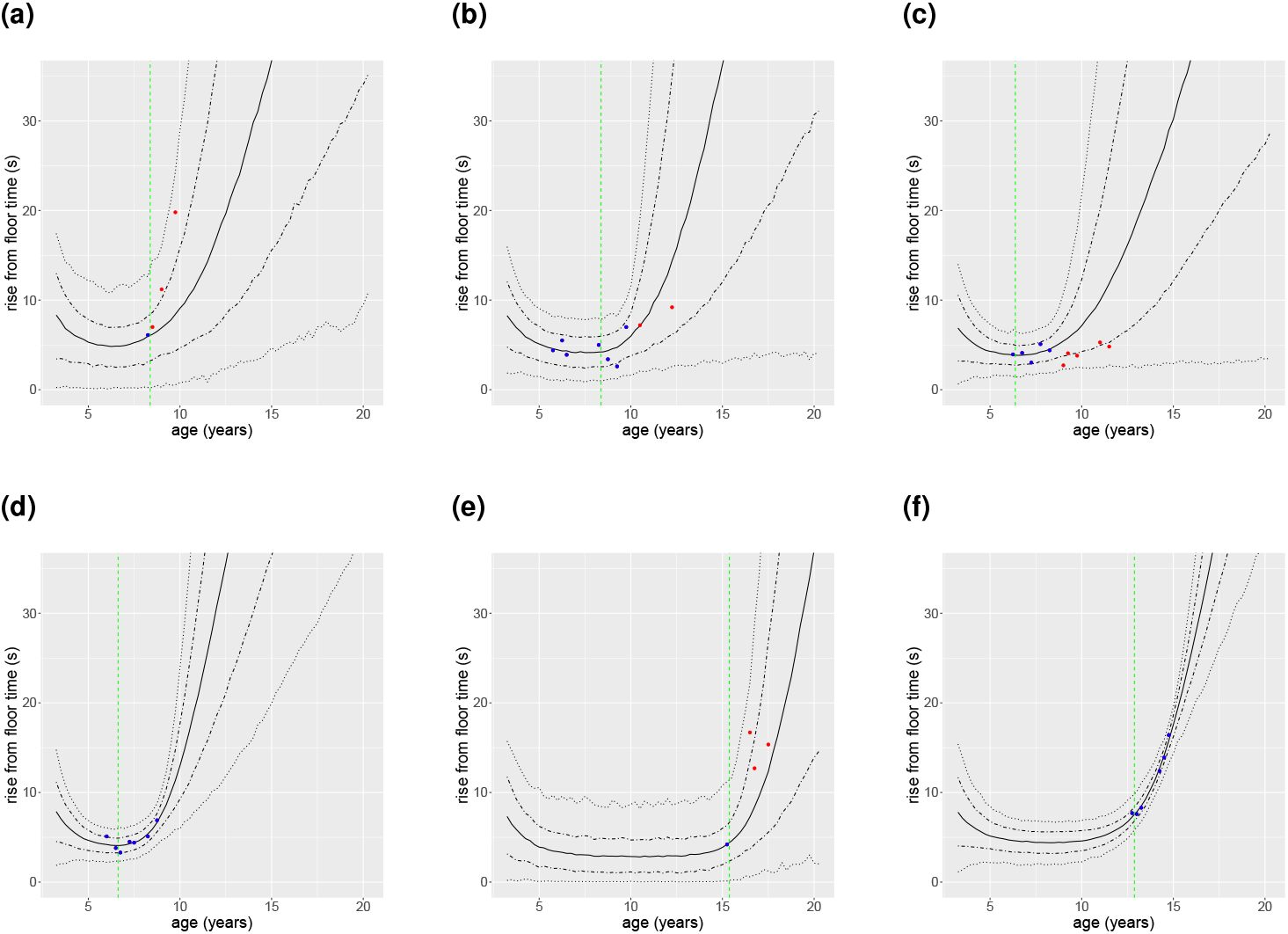
Predictions of rise from floor time for six patient. Blue dots are measurements that are provided to the model, red dots are measurements that were held-aside and were not given to the model. The central black lines represent the median prediction, with the outer lines representing the 70% and 95% prediction intervals. The horizontal green lines represent when a steroid intervention occurred.

These predictions reflect well on the model, indicating good predictive capability. The general pattern of NSAA trajectories seem to be present in each case, but each prediction is also tailored to the specific patient in question. The degree of uncertainty clearly depends on how much data is available for a specific patient, with Figure 2a exhibiting a large amount of uncertainty, a result of only having one measurement of NSAA. Nonetheless, a general decline is still predicted, due to the information borrowed from other patients about how the disease and NSAA progresses. Similarly, the predictions forward in Figure 2f are very concentrated, a result of the greater number of measurements on that patient. This increased confidence might also be because data is available for this patient after a decline has already been observed, and so the model no longer needs to predict when this will occur.

The model is also flexible enough to adapt to different clinical behaviours, with the very late decline of Figure 2e still captured.

The model is not always perfect, as we can see in Figure 2c, where the decline is sooner and sharper than expected. This is not particularly unexpected, as in this case, the patient only declines *just* after the last measurement the model is given. Nonetheless, the 95% prediction interval still almost covers the realised trajectory, indicating that the model acknowledged the extreme observed outcome was a real possibility.

Of course, these predictions are not only for NSAA, but are paired with predictions for walk time and rise from floor time. These predictive plots can be found in the appendix. Additionally, it is worth noting that the ‘spikiness’ of these plots are a result of the Monte Carlo error from estimating the plotted quantiles from a limited number of MCMC samples.

Collectively, the testing data lies within the model’s 95% prediction intervals 95.7% of the time (1573 out of 1644 total validation points). Breaking this down to the individual clinical output predictions, the NSAA testing data lies within the model’s 95% prediction intervals 95.5% of the time (634 out of 673), the walk time data lies within the 95% prediction intervals 94.9% of the time (517 out of 531), and the rise from floor time 96.8% (429 out of 443). Overall, this suggests the model has good predictive accuracy. The rise from floor time predictions are somewhat under-confident, and potentially the NSAA predictions as well, but in general these predictions appear accurate.

It is worth noting that the plots given here provide a visual summary of the predictions a patient can receive, but other useful summaries can be easily extracted for a patient. These summaries might include predictions such as “what is the probability that the patient’s NSAA score drops below 10 before age 12?” or “is this patient expected to perform better than average at age 15”. Key summaries such as these might be easier to communicate to patients and their families, but are just as easily obtained from the model as the trajectory plots.

As a specific note for DMD, the treatment effects of steroids included here appear to have a small effect on a patient’s progression. The mean beneficial effect of being on “daily” steroids (i.e the mean value for any corresponding potential **Γ**_*i*_ entry) is 0.0177, for “intermittent 10.10” this is 0.0004, “other” 0.0074, and “unknown” 0.0032. When compared to a mean value for the disease decline (i.e. the mean estimate for any potential Δ_*i*_ value) of 0.2724, the general impact of the treatments is quite small. This is an improvement on Goemans et al. ^28^, where steroids are found to have a negative effect, but it is nonetheless clear that discerning the true effect of steroids is difficult in observational studies like this. It is reassuring, however, that “daily” steroids are found to have a much greater benefit than “intermittent 10.10” steroids here, as one might expect ^29^. As such, the model is learning that steroids have a positive effect, but it can only distinguish a slight positive effect from the natural variation in patients using the current dataset. The model should have the potential to learn the impact of treatments, and so this small estimate is likely due to a limited amount of varied treatment regime data compared to the variation present in patient outcome, which is a limitation of working with real-world / natural history data (rather than data from a clinical trial).

## Discussion

The model framework outlined here appears capable. It has the ability to predict forward a patient’s trajectory, including the epistemic and aleatoric uncertainty in these predictions; it can tailor the structure of these predictions depending on the features of a specific patient’s clinical history while still borrowing strength from the information in the wider dataset; it has the ability to incorporate the effects of interventions; and it can do so while seamlessly accounting for missing data.

We have presented some preliminary results applying this model to Duchenne Muscular Dystrophy. The predictive performance of this model is good, despite the limited data and wide variation in patient outcome. These predictions have can provide information about how a *specific* patient is likely to progress, which is valuable given the wide spread of experiences different patients can have.

Whilst we did find some benefit to prescribing steroids, the impact of this is smaller than we would have expected. This is likely due to the limitations of the observational dataset we have, where the variety in when steroids are prescribed is quite low (for example, most patients are prescribed steroids just as they begin to decline, which makes it difficult to determine if a slow decline is a result of the steroids, or because the patient naturally declines slowly). Furthermore, many patients are seemingly to be prescribed steroids just *before* their decline (for example, the patient in Figure 2d). This could be because they failed to reach expected developmental milestones, because a decline was observed in other clinical outcomes, or because of evolution of standards of care and their implementation. A more varied dataset, perhaps one that includes healthy individuals as well, allowing for better estimation of the healthy state parameters, and one that includes all relevant clinical outcome measurements, would likely improve the model results.

This model framework could be applied to many other degenerative diseases, including diseases which manifest later in life, due to the inclusion of parameters which control the delay of symptom onset. In some cases, it might be valuable to include an additional additive effect in Equation (1) if it is believed that the treatments can also provide an overall immediate (or with some lag) boost to a patient’s health, rather than only affecting the change over time, as we have assumed for DMD.

Other improvements to our framework can be imagined by making the innovations (the random walk 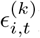) multivariate, or the observational error variance 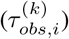 or innovation variance 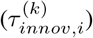 terms themselves multivariate, but that this would be computationally taxing, and is unlikely to have a substantial impact. Furthermore, for even more complex diseases, additional latent disease states could be included, allowing for more complex disease progression to occur. Alternatively, additional 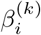 terms could be included, allowing for polynomial responses to the latent disease state to be modelled.

Another direction for future improvement might revolve around computational time. Currently, to generate predictions for a new patient, the model needs to be re-fit using the data from all available patients, which is taxing (although, with long-term chronic degenerative diseases, this is not an insurmountable cost). Because all the information from the other patients is summarised in the group level parameters, such as ***μ***_*α*_, this process could be streamlined by extracting these posterior distributions from a previous fit, and using them as priors for the new patient, avoiding the need to include data from all patients. How to do this remains future work.

Overall, the model framework presented here is a powerful tool, allowing predictions to truly be made patient-specifc even when data on that patient is limited. This can then supplement the expertise of clinicians and aid in their prognosis and treatment of degenerative diseases, and could also assist in the interpretation of real world data efficacy of novel therapies.

## Data Availability

Data may be available upon reasonable request to either Ian Bamsey (; Managing Director at Certus Technology, which hosts the database), or Francesco Muntoni

## Acknowledgements

The authors are grateful to patients for participating in the clinical assessments and agreeing to make their data available for research. The authors would like to acknowledge investigators and staff involved in the UK North Star which is funded by Muscular Dystrophy UK.

North Star UK:

F. Muntoni, A.Y. Manzur, S. Robb, R. Quinlivan, A. Sarkozy, P. Munot, M. Main, L.E. Abbot, H. Patel, S. Samsuddin, V. Ayyar-Gupta, K. Bushby, V. Straub, M. Guglieri, C. Bertolli, A. Mayhew, R. Muni-Lofra, M. James, D. Moat, J. Sodhi, H. Roper, D. Parasuraman, H. McMurchie, R.M. Rabb, A. Childs, K. Pysden, L. Pallant, S. Spinty, G. Peachey, R. Madhu, A.J. Shillington, E. Wraige, H. Jungbluth, V. Gowda, J. Sheehan, R. Spahr, I. Hughes, E. Bateman, C. Cammiss, T. Willis, L. Groves, N. Emery, P. Baxter, M.T. Ong, N. Goulborne, M. Senior, E. Scott, L. Hartley, L.B. Parsons, A. Majumdar, K. Vijaykumar, F.F. Mason, L. Jenkins, B. Toms, C.H. Frimpong-Ansah, K. Naismith, J. Dalgleish, A. Keddie, I. Horrocks, M. Di Marco, J. Dunne, G.C.S. Chow, A. Miah, C. de Goede, A. Selley, N. Thomas, M. Illingworth, M. Geary, J. Palmer, C.P. White, K. Greenfield, S. Tiraputhi, S. MacAuley, N. Hussain, H. Robbins, M. Iqbal, G. Ambegaonkar, D. Krishnakumar, C. Ward, J. Taylor, A. O’Hara, J. Tewnion, S.R. Chandratre, S. Ramdas, M. White, H. Ramjattan, A. Baxter, J. Yirrel

## Funding

EB, KTA, and PC gratefully acknowledge the financial support of the EPSRC via grant EP/T017856/1. FM is partially supported by the National Institute for Health Research (NIHR) Biomedical Research centre at Great Ormond Street Hospital for Children NHS Foundation Trust and University College London, as well as by Muscular Dystrophy UK.

## Additional Results plots

Plots of example model predictions for NSAA were provided in Section. Here, these same predictions are provided but for walk time and rise from floor time.

## Notes

### Competing Interest Statement

The authors have declared no competing interest.

### Funding Statement

This work received funding from the EPSRC via grant
EP/T017856/1. Support was also provided by the National Institute for Health Research (NIHR)
Biomedical Research centre at Great Ormond Street Hospital for Children NHS Foundation Trust
and University College London, as well as by Muscular Dystrophy UK.

### Author Declarations

This work used data obtained previously by the NorthStarUK network, and approval was given to perform this work. The North Star clinical network study was reviewed by the North Sheffield Research Ethics Committee and it was felt that it did not need to go to a research ethics committee. Written informed consent was obtained for the collection of all clinical data and the NorthStar clinical network project has Caldicott Guardian approval. All clinical assessments are conducted according to the principles of the Declaration of Helsinki (2000) and its later amendments and the Principles of Good Clinical Practice.

